# Development and Validation of a Computable Phenotype for Turner Syndrome Utilizing Electronic Health Records from a National Pediatric Network

**DOI:** 10.1101/2023.07.19.23292889

**Authors:** Sarah D. Huang, Vaneeta Bamba, Samantha Bothwell, Patricia Y. Fechner, Anna Furniss, Chijioke Ikomi, Leena Nahata, Natalie J Nokoff, Laura Pyle, Helina Seyoum, Shanlee M Davis

## Abstract

Turner syndrome (TS) is a genetic condition occurring in ∼1 in 2,000 females characterized by the complete or partial absence of the second sex chromosome. TS research faces similar challenges to many other pediatric rare disease conditions, with homogenous, single-center, underpowered studies. Secondary data analyses utilizing Electronic Health Record (EHR) have the potential to address these limitations, however, an algorithm to accurately identify TS cases in EHR data is needed. We developed a computable phenotype to identify patients with TS using PEDSnet, a pediatric research network. This computable phenotype was validated through chart review; true positives and negatives and false positives and negatives were used to assess accuracy at both primary and external validation sites. The optimal algorithm consisted of the following criteria: female sex, ≥1 outpatient encounter, and ≥3 encounters with a diagnosis code that maps to TS, yielding average sensitivity 0.97, specificity 0.88, and C-statistic 0.93 across all sites. The accuracy of any estradiol prescriptions yielded an average C-statistic of 0.91 across sites and 0.80 for transdermal and oral formulations separately. PEDSnet and computable phenotyping are powerful tools in providing large, diverse samples to pragmatically study rare pediatric conditions like TS.

## INTRODUCTION

Affecting about 1 in 2,000 live female births, Turner syndrome (TS) is a rare genetic condition characterized by the complete or partial absence of the second sex chromosome (Berglund et al., 2020). Individuals with TS experience a wide variety of clinical manifestations, including cardiac defects, short stature, premature ovarian failure, and neurodevelopmental conditions (Gravholt et al., 2017). Guidelines recommend treatment of premature ovarian failure with transdermal low-dose estradiol starting between 11-12 years of age to promote timely physical and social development (Gravholt et al., 2017). As with other sex chromosome aneuploidies and pediatric rare conditions in general, our knowledge of TS is largely based on small single-center studies challenged by multiple biases. With cell free DNA (cfDNA) screening increasing prenatal identification, there is an ever-growing need for generalizable research that informs best clinical practice and improves health outcomes for individuals with TS (Bussolaro et al., 2023).

Electronic Health Record (EHR) data analytics is a powerful tool that can address many of the limitations associated with past pediatric TS research. PEDSnet is an innovative national network that uses a common data model to combine EHR for millions of children from eight pediatric institutions across the United States (Forrest et al., 2014). It was created to promote multi-institutional collaboration for pediatric studies, facilitate research that will have direct clinical applications that improve child health, and reduce time and expense of conducting research (Forrest et al., 2014). It is a valuable resource that offers larger, more diverse samples to better pediatric research for all conditions and has the potential to be especially impactful for rare disease conditions like TS. The PEDSnet infrastructure not only supports retrospective analyses of EHR data but also facilitates identification of disease cohorts for comparative outcome studies and pragmatic clinical trials (Forrest et al., 2014; Wenderfer et al., 2022). To prepare for such studies in TS, a computable phenotype, or algorithm that identifies girls with TS in a real world EHR dataset, is needed.

Computable phenotyping uses a defined set of criteria to automate cohort identification (Richesson et al., 2013). Criteria can include various discrete data elements within the EHR, such as billing diagnoses, problem lists, medications, laboratory results, orders, demographics, and more. When applied to a group of patients, it automatically identifies a cohort of interest. Although computable phenotyping guarantees efficiency, it is also crucial to assess accuracy as solely utilizing a single billing diagnosis can lead to poor specificity in cohort identification, especially for genetic conditions (Richesson et al., 2013). Multiple computable phenotypes have been developed and validated, including for specific cohorts of pediatric cancer and Crohn’s disease patients in PEDSnet, however this approach has not been previously used in pediatric genetic conditions like TS (Denburg et al., 2019; Khare et al., 2020; Phillips et al., 2019; Wenderfer et al., 2022). In this study, we aimed to develop and validate a computable phenotype that accurately identifies patients with TS in PEDSnet for use in future studies. Given recent literature has called for comparative effectiveness studies for hormone replacement therapy (HRT), we further aimed to assess the accuracy of estradiol treatment and formulation in this population (Dowlut-McElroy & Shankar, 2023).

## METHODS

### Data Source

Institutions contributing data for this study included Children’s Hospital Colorado (CHCO, primary site), Children’s Hospital of Philadelphia (CHOP), Seattle Children’s Hospital, Nemours Children’s Health, and Nationwide Children’s Hospital. Deidentified patient-level data were securely obtained from the PEDSnet Data Coordinating Center for all individuals meeting the initial algorithm criteria (described below), in addition to a random sample of individuals meeting all algorithm criteria except a TS diagnosis. Records were re-identified locally to allow for chart review, and de-identified data were collected and managed using REDCap (Research Electronic Data Capture) tools hosted at the University of Colorado (Harris et al., 2009). Analyses were performed in R Statistical Software (v4.1.2; R Core Team 2021).

### TS Computable Phenotype Development

An initial algorithm was developed using clinician and investigator experience, as well as incorporation of similar efforts utilizing PEDSnet in studies of pediatric cancer and gastrointestinal conditions (Khare et al., 2020; Phillips et al., 2022). The initial algorithm consisted of the following criteria to identify patients with TS who were potentially eligible for HRT: female sex, ≥1 outpatient encounter between 2009-2019, ≥10 years old at the time of the most recent encounter, and ≥1 diagnosis code in their billing diagnoses and/or problem list mapping to TS (Supplemental Table 1). The algorithm was tested at one institution (primary analytic cohort), with the plan to evaluate false positives and negatives to further refine criteria to include encounter specialties, medications, or growth parameters if appropriate.

### Analytic Cohorts

The primary analytic cohort included 306 individuals meeting inclusion criteria regardless of phenotypic severity at CHCO (286 with TS diagnosis and 20 without). Blinded research assistants at CHCO who were specifically trained in the necessary data elements (SH; HS) reviewed individual EHRs for all subjects and used the standardized REDCap case report form to record whether the individual had a genetically confirmed diagnosis of TS, as well as the diagnosis date, karyotype, HRT status, HRT formulation, and HRT start date. If the diagnosis of TS or HRT status were unclear, a board-certified pediatric endocrinologist with expertise in TS (SD) reviewed the chart to make a final determination. To test the external validity of the most optimal algorithm TS diagnosis codes, random samples of 40 patients (30 meeting inclusion criteria and 10 meeting all criteria except TS diagnosis) were selected at the four other PEDSnet institutions. These individuals were reidentified for chart review with the same REDCap case report form. Investigators carrying out the chart reviews at each of these institutions were also board-certified pediatric endocrinologists familiar with treating TS and blinded to the PEDSnet diagnosis status.

### Primary Analysis

The primary outcome was a confirmed diagnosis of TS. True positives and negatives and false positives and negatives were used to calculate sensitivity, specificity, positive predictive value (PPV), negative predictive value (NPV), accuracy, and concordance statistic (C-statistic, or the probability patients with TS have a higher risk score than patients without) for the primary outcome. False positive and negative cases were individually reviewed by a second investigator to determine the reason for the incorrect classification and allow refinement of the original algorithm. Beyond the original criteria of ≥1 diagnosis code mapping to TS, criteria of ≥2, ≥3, ≥4, and ≥5 diagnoses codes also mapping to TS were applied to the primary analytic sample while all other components of the algorithm remained the same. Sensitivity, specificity, PPV, NPV, accuracy, and C-statistic were calculated for each of these algorithms and the algorithm with the highest C-statistic using the CHCO cohort was selected as the optimal algorithm. This optimal algorithm was then tested in the validation cohort comprised of the four other sites.

### Analyses of Secondary Outcomes

To determine the accuracy of the age of first TS diagnosis coded in PEDSnet, the difference between PEDSnet and chart review determined age of diagnosis was determined for each individual. The mean individual difference and range were then compared between PEDSnet and chart review for CHCO only, other sites only, and all sites combined using two-sided t-tests with a p-value <0.05 considered significant.

To determine how well accurately we could determine a patient’s HRT status (yes or no; Supplemental Table 1) using PEDSnet data, true positives and negatives and false positives and negatives were used to calculate sensitivity, specificity, PPV, NPV, accuracy, and C-statistic at all sites. Since the data were obtained from PEDSnet in 2019 but not all data for 2019 were available, a sensitivity analysis was run to exclude patients who had started HRT in 2019 and the results were similar. The same approach was used to determine the estradiol formulation (transdermal and oral), with the assumption that all estradiol prescriptions not specifying a formulation (e.g. “estrogens”) were oral route. Finally, the accuracy of the age of HRT initiation was tested using a two-sided t-test comparing the mean individual difference between PEDSnet and chart review for age of first estradiol prescription.

### Editorial Policies and Considerations

This study was approved by the institutional review board (IRB) at CHOP (#14–011242, 16– 012878, and 16–013563) with a waiver of consent. A Master Reliance Agreement was in place between all institutions involved for CHOP to be the single IRB overseeing computable phenotyping within PEDSnet. Re-identification took place at the local sites and only de-identified data were shared with CHCO for conducting the analyses.

## RESULTS

### Sample Description

Demographics for the TS cases and non-cases obtained from PEDSnet are shown in Table 1. The cases and non-cases were of similar age, duration in PEDSnet, race, ethnicity, and insurance status and generally representative of a clinical pediatric cohort in the US. Across sites, there were on average 9.0 years of EHR data per patient. There were two cases of indeterminant TS diagnosis due to low level monosomy X mosaicism (45,X in 1 of 50 cells analyzed) and additional genetic testing was not available. These two cases were excluded from the analysis.

**Table 1:**
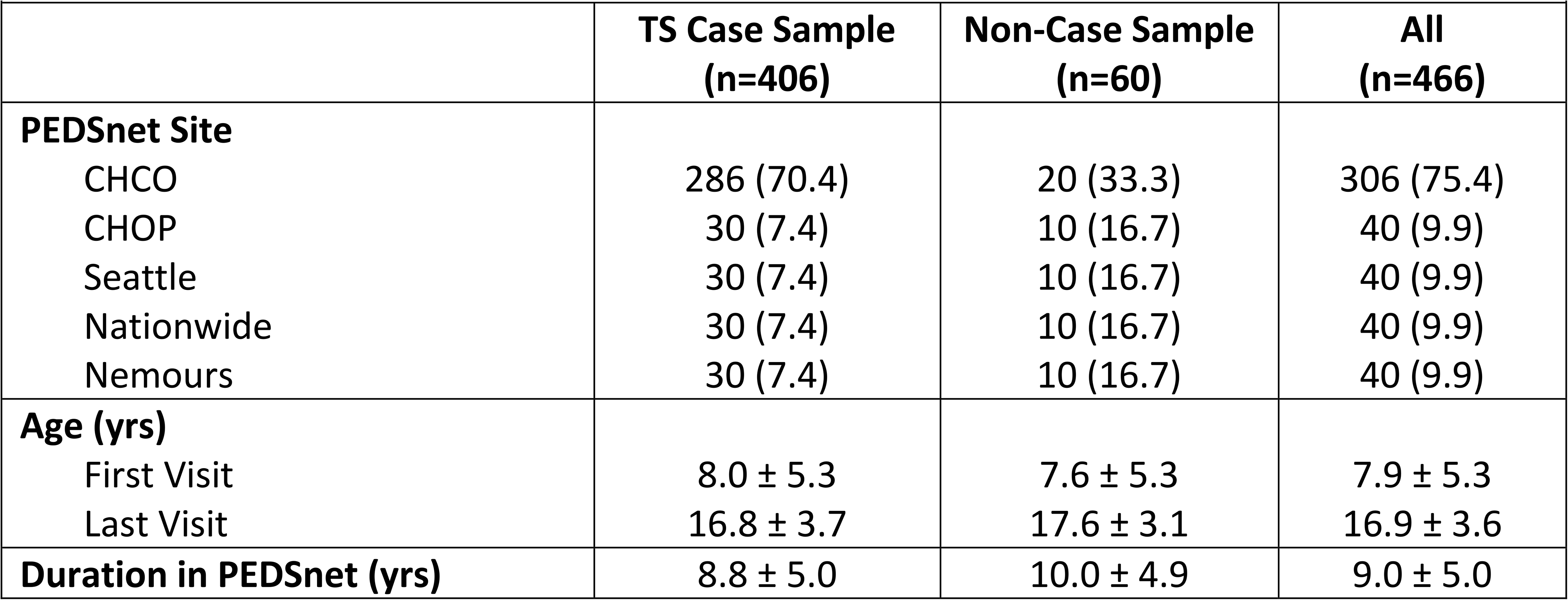
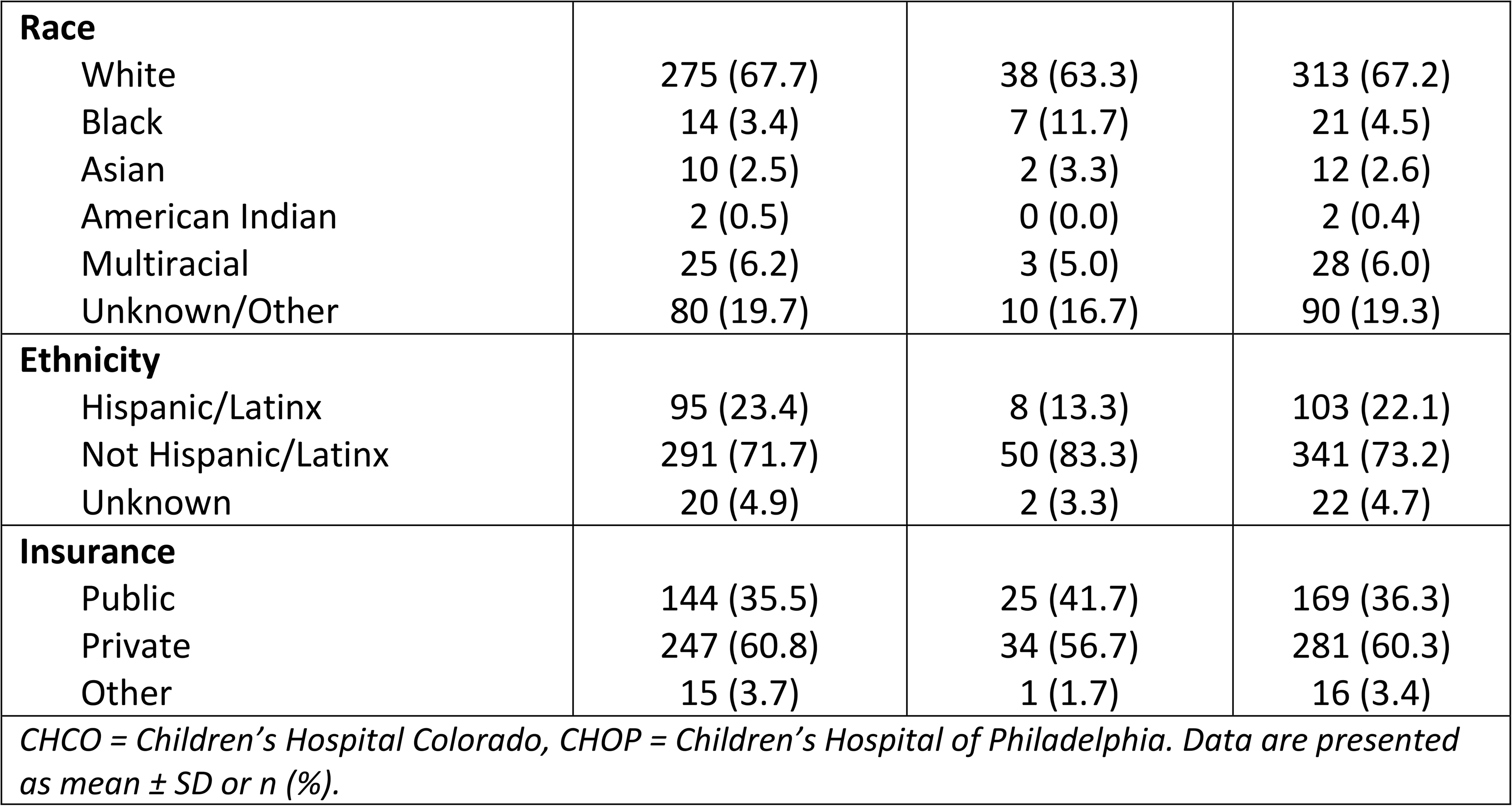
Sample description and demographics from PEDSnet.

### Classification Accuracy

The initial algorithm applied at CHCO identified a total of 14 false positives yielding a suboptimal C-statistic of 0.79. Of the false positives, 100% had a common TS characteristic, 92.9% had undergone genetic testing, and 64.3% had been diagnosed with a condition under a larger umbrella such as a different genetic or difference in sex development condition (Supplemental Table 2). Given these findings, the addition of other features that characterize TS to the computable phenotype (i.e. short stature, delayed puberty, premature ovarian failure) did not yield a higher C-statistic; neither did the addition of clinical encounter specialties (i.e. endocrine, genetics) or medication/test orders (growth hormone, estradiol prescription, genetic testing, etc.) as the number of false negatives greatly increased with minimal increase in false positives. However, increasing the number of TS diagnosis codes within the EHR was able to improve the specificity without significantly compromising sensitivity, resulting in a higher C-statistic (Table 2). The algorithm with 3 TS billing diagnoses yielded the highest C-statistic of 0.92 as well as the best balance of sensitivity and specificity, with values of 93.4% and 91.2% respectively, and was therefore identified as the most optimal (Figure 2).

**Figure 1:**
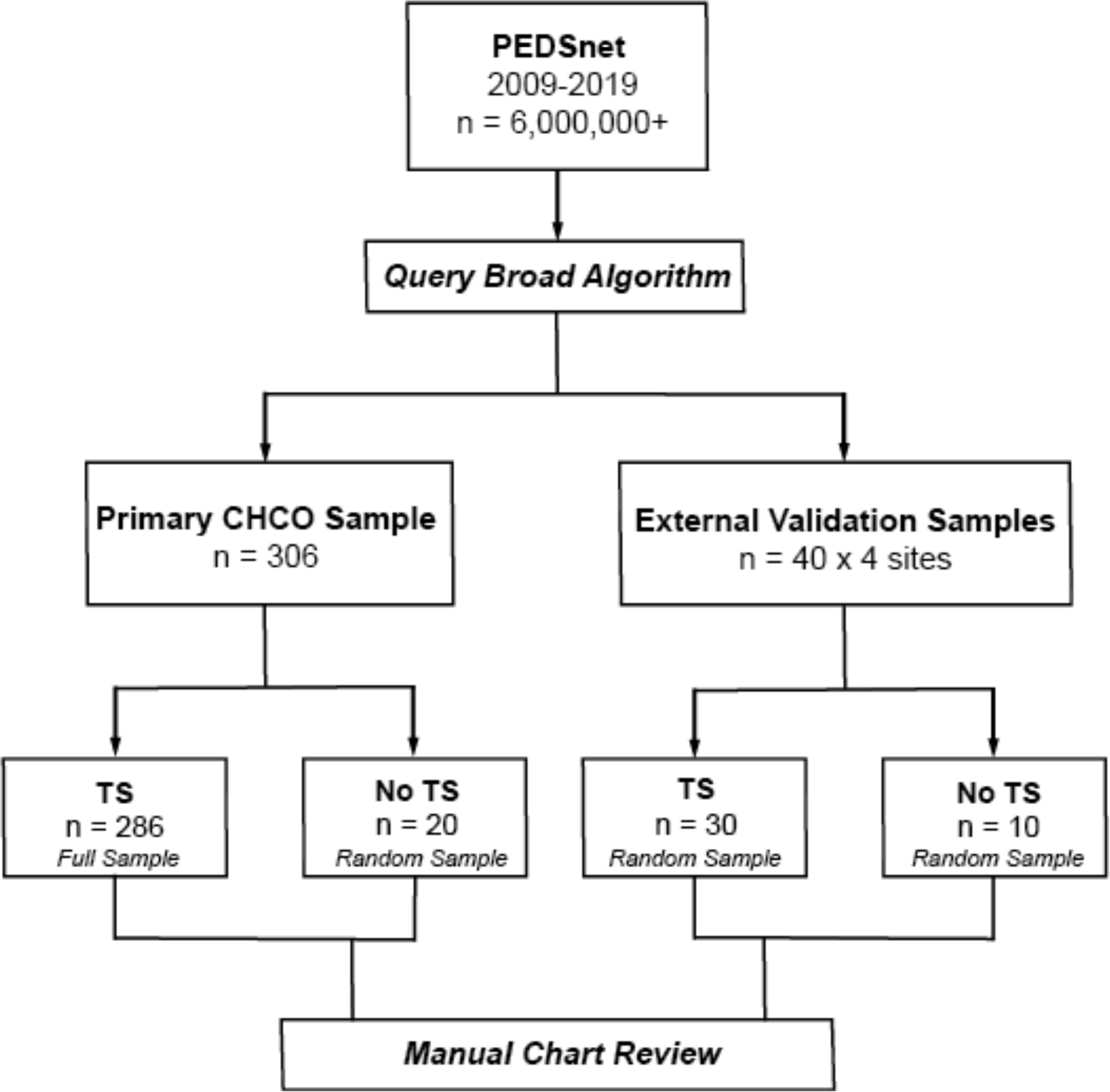
Flow diagram for cohort identification and algorithm development. A broad algorithm was applied to PEDSnet data to identify the primary sample for algorithm development and the validation sample at four additional sites. Samples consisted of patients diagnosed with TS and patients without TS per PEDSnet. Manual chart review was conducted for 466 unique patients.

**Figure 2:**
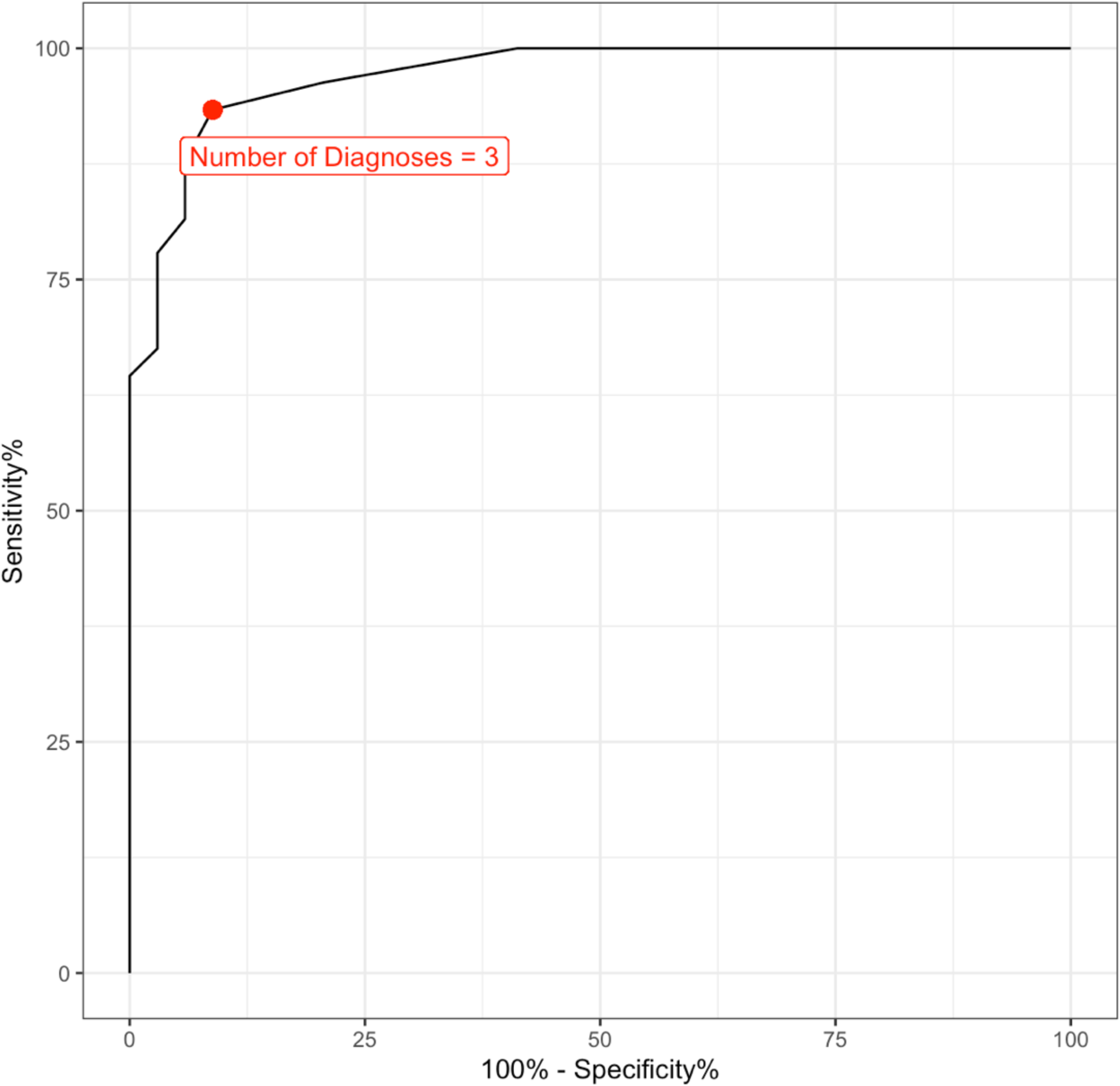
ROC Curve Analysis to determine the optimal number of Turner syndrome diagnoses. Visualization of the sensitivity (y axis) and specificity (x-axis) based on the number of TS diagnoses.

**Table 2:** Applying varying algorithms to the primary CHCO sample.

| **Table 2: Applying varying algorithms to the primary CHCO sample.** |  |  |  |  |  |  |  |  |  |  |
| --- | --- | --- | --- | --- | --- | --- | --- | --- | --- | --- |
| Algorithm (# of TS diagnoses) | True Positives (TP), n | False Positives (FP), n | True Negatives (TN), n | False Negatives (FN), n | Sensitivity % (95% CI) | Specificity % (95% CI) | PPV % | NPV % | Accuracy % (95% CI) | C-statistic |
| 1 | 271 | 14 | 20 | 0 | 100 (98.7-100) | 58.8 (40.7-75.4) | 95.1 (92.6-97.6) | 100 (100-100) | 95.4 (92.4-97.5) | 0.79 (0.71-0.88) |
| 2 | 261 | 7 | 27 | 10 | 96.3 (93.3-98.2) | 79.4 (62.1-91.3) | 97.4 (95.5-99.3) | 73.0 (58.7-87.3) | 94.4 (91.2-96.7) | 0.88 (0.81-0.94) |
| 3 | 253 | 3 | 31 | 18 | 93.4 (89.7-96.0) | 91.2 (76.3-98.1) | 98.8 (97.5-100) | 63.3 (49.8-76.8) | 93.1 (89.7-95.7) | 0.92 (0.87-0.97) |
| 4 | 238 | 2 | 32 | 33 | 87.8 (83.3-91.5) | 94.1 (80.3-99.3) | 99.2 (98.1-100) | 49.2 (37.0-61.4) | 88.5 (84.4-91.9) | 0.91 (0.86-0.95) |
| 5 | 234 | 2 | 32 | 37 | 86.4 (81.7-90.2) | 94.1 (80.3-99.3) | 99.2 (98.1-100) | 46.4 (34.3-58.5) | 87.2 (82.9-90.1) | 0.90 (0.85-0.94) |
| Data are presented as N or % with 95% confidence interval in parentheses. Excludes one case in which Turner syndrome status could not be determined from chart review. |  |  |  |  |  |  |  |  |  |  |

Table 3 details the results when the most optimal 3-diagnosis algorithm was applied to an external validation sample at other institutions. Similar to the CHCO sample, the algorithm produced a reasonable balance of sensitivity and specificity across all sites, and all C-statistic values were >0.89. The average sensitivity across all sites was 97.0%, specificity 87.9%, PPV 93.6%, NPV 90.0%, accuracy 93.6%, and C-statistic 0.93.

**Table 3:**
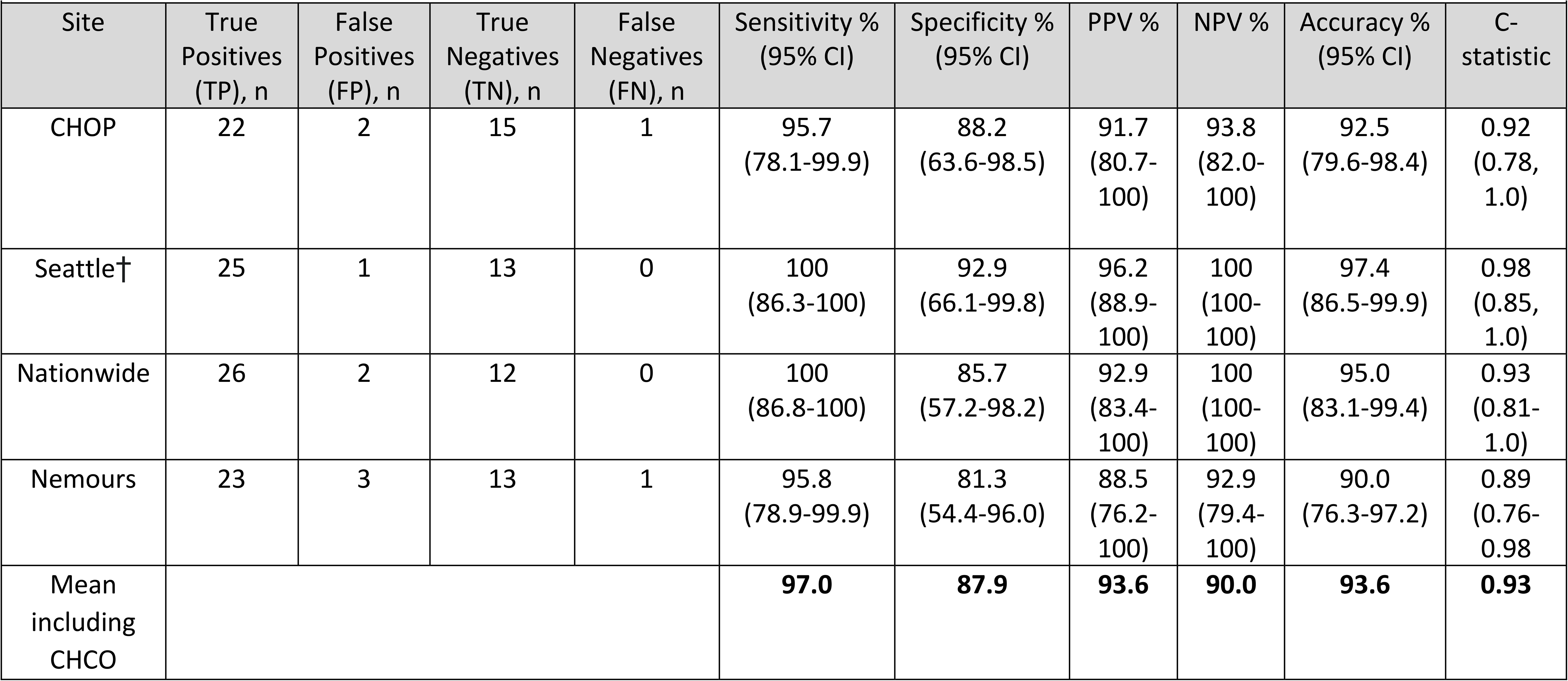
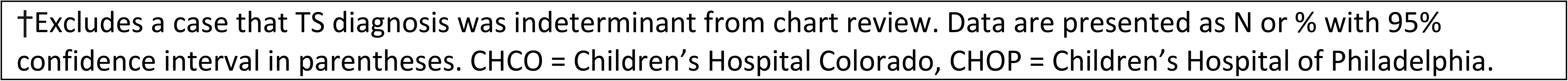
Applying the 3-diagnosis algorithm for TS diagnoses to external validation samples.

|  |  |  |  |  |  |  |  |  |  |  |
| --- | --- | --- | --- | --- | --- | --- | --- | --- | --- | --- |
| Site | True Positives (TP), n | False Positives (FP), n | True Negatives (TN), n | False Negatives (FN), n | Sensitivity % (95% CI) | Specificity % (95% CI) | PPV % | NPV % | Accuracy % (95% CI) | C-statistic |
| CHOP | 22 | 2 | 15 | 1 | 95.7 (78.1-99.9) | 88.2 (63.6-98.5) | 91.7 (80.7-100) | 93.8 (82.0-100) | 92.5 (79.6-98.4) | 0.92 (0.78, 1.0) |
| Seattle† | 25 | 1 | 13 | 0 | 100 (86.3-100) | 92.9 (66.1-99.8) | 96.2 (88.9-100) | 100 (100-100) | 97.4 (86.5-99.9) | 0.98 (0.85, 1.0) |
| Nationwide | 26 | 2 | 12 | 0 | 100 (86.8-100) | 85.7 (57.2-98.2) | 92.9 (83.4-100) | 100 (100-100) | 95.0 (83.1-99.4) | 0.93 (0.81-1.0) |
| Nemours | 23 | 3 | 13 | 1 | 95.8 (78.9-99.9) | 81.3 (54.4-96.0) | 88.5 (76.2-100) | 92.9 (79.4-100) | 90.0 (76.3-97.2) | 0.89 (0.76-0.98) |
| Mean including CHCO |  |  |  |  | **97.0** | **87.9** | **93.6** | **90.0** | **93.6** | **0.93** |

†Excludes a case that TS diagnosis was indeterminant from chart review. Data are presented as N or % with 95% confidence interval in parentheses. CHCO = Children's Hospital Colorado, CHOP = Children's Hospital of Philadelphia.

### Age of TS Diagnosis

Table 4 compares the age of TS diagnosis per PEDSnet and the chart review at CHCO, other sites, and all sites combined. Using the age of first diagnosis in PEDSnet overestimated the true age of diagnosis by 3.7 years (p<0.001). The mean individual difference range was large (–1.0-18.9), with an age of diagnosis identified in PEDSnet being up to 19 years later than the chart review-identified diagnosis age. The plots displaying these data (Figure 3) further emphasize the greater average TS diagnosis age per PEDSnet compared to the chart review, with many cases diagnosed prenatally or within the first year of life not reported in PEDSnet until much later.

**Figure 3:**
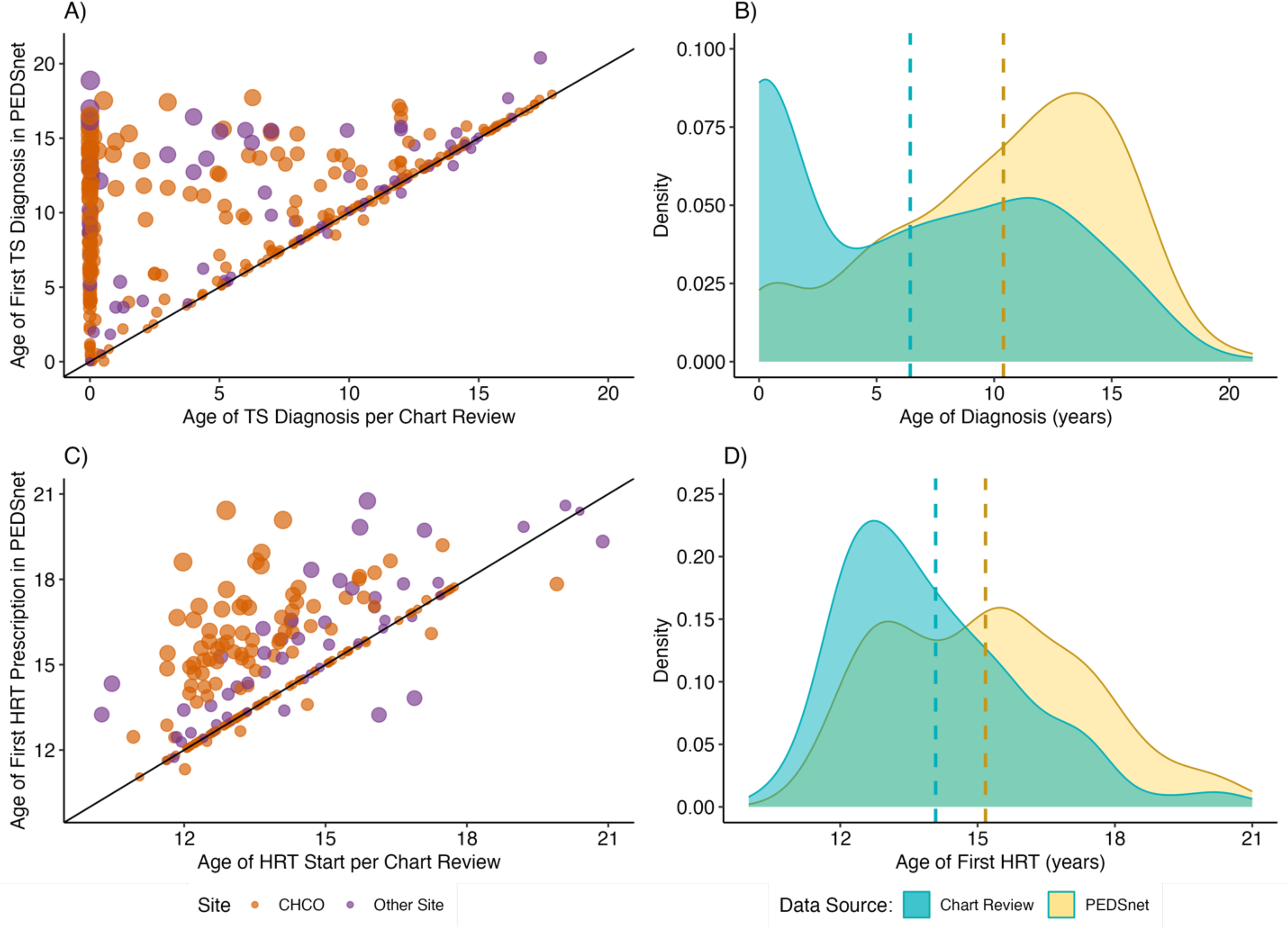
(A) Scatterplot illustrating discrepancy in age of Turner syndrome diagnosis estimated by PEDSnet vs confirmed by chart review. Dots represent individuals and size of the dots represent discrepancy from the regression line. (B) Density plot of the age of TS diagnosis per chart review (blue) and PEDSnet (yellow), with median values for each represented with dashed lines (median difference of 4 years). (C) Scatterplot demonstrating discrepancies in age of estradiol initiation between chart review and PEDSnet. (D) Density plot of the age of which estradiol treatment was initiated per chart review (blue) and PEDSnet (yellow).

**Table 4:** Age of TS diagnosis (in years) estimated in PEDSnet compared to chart review.

|  | **PEDSnet**<br>(mean ± SD) | **Chart Review**<br>(mean ± SD) | **p-value** | **Mean Individual Difference** | **Mean Individual Difference Range** |
| --- | --- | --- | --- | --- | --- |
| **CHCO** | 10.2 ± 5.1 | 6.1 ± 5.8 | p<0.001 | 3.8 ± 4.8 | -1.0-18.0 |
| **Other Sites** | 11.0 ± 4.7 | 7.4 ± 5.5 | p<0.001 | 3.5 ± 4.9 | -0.9-18.9 |
| **All Sites Combined** | 10.4 ± 5.0 | 6.4 ± 5.7 | p<0.001 | 3.7 ± 4.9 | -1.0-18.9 |

Data include cases with a TS diagnosis in PEDSnet and confirmed by chart review (n=375). Data are presented as mean ± SD or as age ranges. CHCO = Children's Hospital Colorado

### HRT Analysis

PEDSnet identified 216/375 (57.6%) of TS cases to have one or more prescriptions for estradiol HRT, whereas 258 (68.8%) were found to have prescriptions for estradiol per chart review. The HRT status was inaccurately classified in 46 (12.3%), meaning PEDSnet’s account of HRT did not accurately reflect these patients’ true HRT status. The average sensitivity across all sites was 90.0%, specificity 96.0%, PPV 97.8%, NPV 81.3%, accuracy 91.2%, and C-statistic 0.93 (Table 5). There was considerable variability between sites for accuracy of HRT formulation. Calculations could not be done for Seattle as the estradiol formulation in PEDSnet was non-specific (i.e. “estrogens” or “estradiol”). The C-statistic for the other sites ranged from 0.75-0.91 (Table 5). The average age of first estradiol prescription was overestimated in PEDSnet by 1.2 years at 15.3 years compared to 14.1 years found in chart review (Table 6).

**Table 5.**
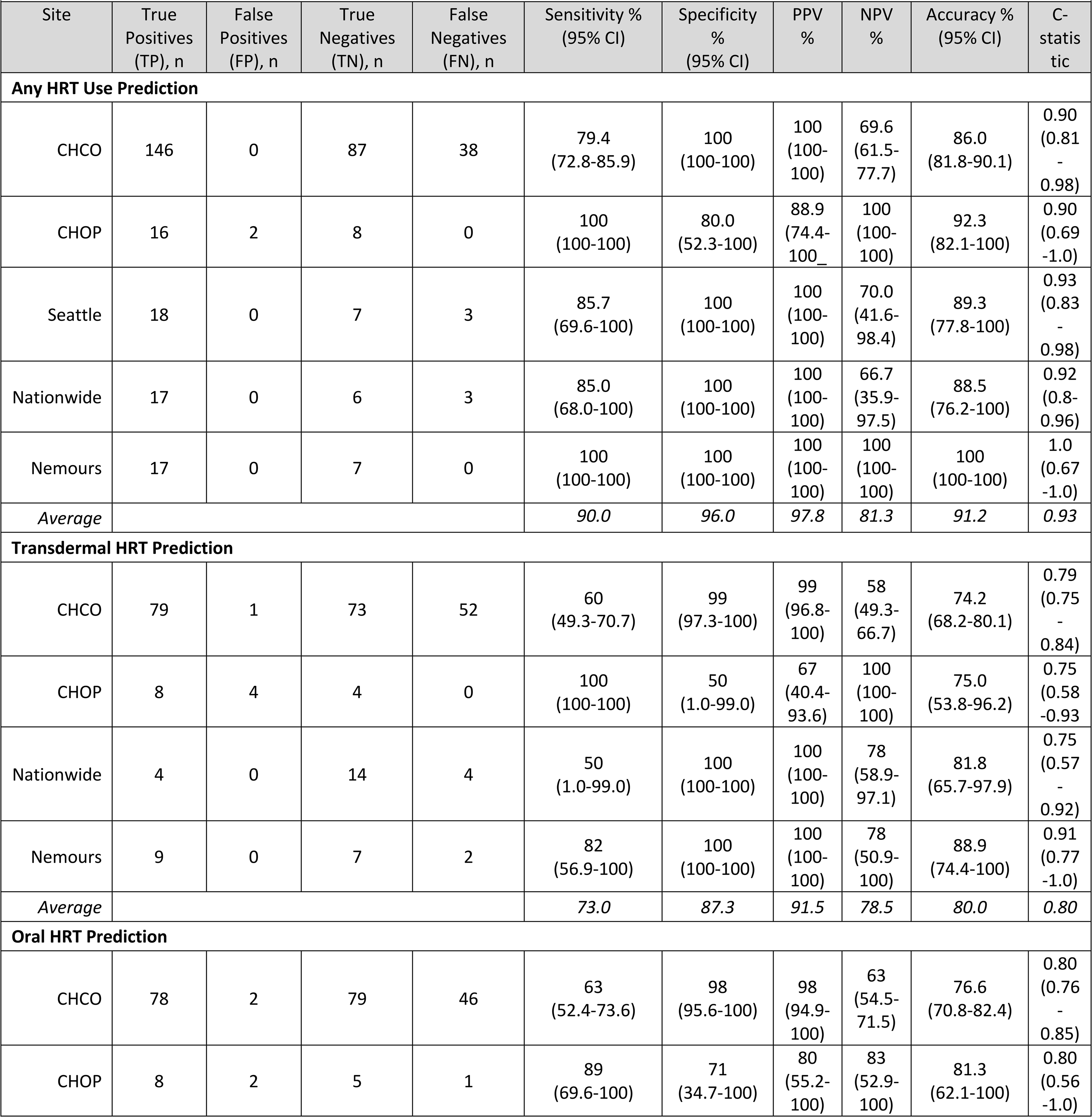
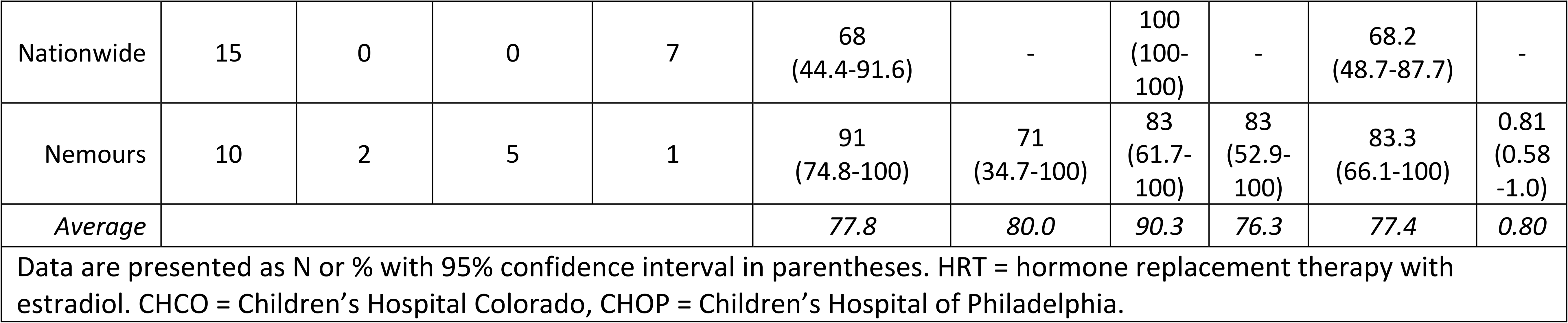
HRT exposure in PEDSnet compared to chart review.

**Table 6:** Accuracy of age of HRT initiation (in years) estimated in PEDSnet compared to chart review.

|  |  |  |  |  |  |
| --- | --- | --- | --- | --- | --- |
|  | PEDSnet<br>(mean ± SD) | Chart Review<br>(mean ± SD) | p-value | Mean Individual<br>Difference | Mean Individual<br>Difference Range |
| **CHCO** | 15.2 ± 2.2 | 13.9 ± 1.9 | p<0.001 | 1.3 ± 1.7 | -2.1-7.5 |
| **Other Sites** | 15.6 ± 2.8 | 14.6 ± 2.3 | p<0.001 | 1.0 ± 1.8 | -3.1-7.0 |
| **All Sites**<br>**Combined** | 15.3 ± 2.4 | 14.1 ± 2.1 | p<0.001 | 1.2 ± 1.7 | -3.1-7.5 |
| HRT = hormone replacement therapy with estradiol.<br>CHCO = Children's Hospital Colorado. Data are presented as mean ± SD or as age ranges. |  |  |  |  |  |

## DISCUSSION

This study aimed to use EHR data to develop and validate a computable phenotype for pediatric patients with TS to facilitate future studies for patients with this condition. Using a minimum of three separate occurrences of a TS diagnosis produced the optimal algorithm with the greatest balance of sensitivity and specificity, yielding an average C-statistic of 0.93 across all sites (values >0.9 are generally considered strong models with high probability in correctly identifying group members) (Westreich et al., 2011). This computable phenotype algorithm accurately identifies pediatric TS cases from EHR data and can be adapted to maximize sensitivity or specificity depending on the study priorities. While the determination of HRT exposure was also acceptable, prediction of the age of TS diagnosis, HRT formulation, and age of HRT initiation were less optimal. These results establish a foundation for identifying TS cases in EHR databases that can be applied in future studies and potentially expanded to other pediatric rare diseases and/or genetic conditions.

Utilizing EHR datasets to study rare conditions in pediatric populations has undeniable benefits. Classic clinical trials require vast resources to achieve adequate sample sizes and are also subject to ascertainment biases that make generalizability to broader clinic populations questionable. Large health networks, such as PEDSnet, can overcome these limitations but rely on EHR data that inevitably contain errors. Relying solely on a single billing code can result in poor specificity, emphasizing the importance of developing and validating a disease-specific computable phenotype algorithm. While computable phenotypes have been developed for several pediatric conditions, to our knowledge this is the first study to undertake this challenge for a genetic condition. As expected, we found that higher sensitivity was achieved with fewer TS billing diagnoses, whereas higher specificity was achieved with a greater number of TS billing diagnoses, with the optimal minimum number of three or more (Supplemental Table 1). Alternative algorithm metrics such as type of specialty encounters, features common in TS, or treatments used in TS did not improve the accuracy of the computable phenotype algorithm as most false positives were due to “rule out TS” cases, which was also identified in Khare et al.’s Crohn’s disease study (2020). Furthermore, requiring these metrics within the algorithm would reduce the ability to accurately study some TS outcomes, such as phenotypic variability, healthcare utilization within specific specialties, and interventions. Importantly, this TS computable phenotype algorithm can and should be adjusted depending on the goals of the specific study. For example, a study utilizing a TS computable algorithm to identify potentially eligible patients that will later be confirmed may wish to maximize sensitivity, whereas a study evaluating outcomes from an intervention relying on EHR data alone may need to maximize specificity for the TS diagnosis. Achieving the best balance of sensitivity and specificity, our three-diagnosis algorithm performed well across multiple institutions and can be considered the preferred TS computable phenotype in the absence of project-specific justification.

Not surprisingly, the age of TS diagnosis is overestimated by PEDSnet data. The date of full Epic implementation at PEDSnet institutions varies, and many patients with TS initially receive a diagnosis by their pediatrician and are later referred to specialty clinics, contributing to a small and predictable delay between actual diagnosis and billing diagnosis within the PEDSnet system. However, as identified by chart review, some patients transfer care into the PEDSnet system after having received care elsewhere for an undefined time period; unfortunately, this delay cannot be reliably accounted for and likely contributes to the large individual variability (–1.0 to +18.9 years). Converting genetic test results into discrete data elements in the EHR or flowsheets allowing clinicians to enter the date of TS diagnosis could facilitate more accurate data; however they are not currently standard among EHR systems or PEDSnet common data model at this time.

Determining who received HRT, and by extrapolation experienced ovarian failure, had good specificity but lower sensitivity. The relatively larger number of false negatives for HRT status may have been present for a variety of reasons. HRT may have been prescribed before the PEDSnet common data structure for medications was developed or implemented by individual institutions. Additionally, HRT prescriptions may have been included in free-text data (e.g. chart notes, etc.) but not entered as a discrete data element, an indication outside medications were not reconciled in the EHR. Unfortunately, determination of HRT formulation was more challenging and varied between sites; we did not attempt to assess dose as this specificity was often not available in the PEDSnet database for estradiol. Future initiatives, such as using a variable selection strategy or random forest modeling to identify top predictors, could explore more variables that may better predict accuracy for HRT. Although we did not specifically examine other interventions, such as growth hormone, we would anticipate similar challenges with the current data structure. TS studies dependent on accurate classification of medications would need to narrow the study population to reduce errors. For example, limiting the sample to individuals who have had multiple visits within endocrinology and/or gynecology would likely improve the accuracy of HRT prescription data.

When considering the utility of PEDSnet or similar data sources to study clinical outcomes in TS, it is also important to recognize that variables that do not currently exist as discrete data elements within the EHR cannot be accessed. Similarly, variables that may exist as discrete data elements but are not part of the PEDSnet common data model are also unavailable. Therefore, scientific questions that require variables such as specific karyotypes, timing of thelarche and menarche, and bone age results would not be feasible within PEDSnet at this time. Developing EHR forms to capture TS-relevant data elements could address these limitations, however this approach would need to be broadly adapted into standard clinical practice in order to be successful.

Validation of the TS computable phenotype across multiple institutions with systematic individual chart reviews is a strength of the current study. However, we only utilized one data source (PEDSnet) with institutions that primarily use the same EHR (Epic), therefore we cannot confirm that the high accuracy of this computable phenotype is translatable to other data sources. This limitation is particularly important if attempting to apply our computable phenotype in an adult obstetrical population where a person pregnant with a fetus affected by TS may be coded as TS in encounter diagnoses. Second, our decision to review a disproportionately smaller number of non-TS controls artificially inflates the sensitivity of our initial algorithm. However, given the rarity of TS, it would be unlikely for false negatives to be present in a larger randomly selected sample. Furthermore, NPV may be underrepresented in the TS diagnosis validation given that the proportion of cases were far greater in our test sample than in the general clinic population, also due the rarity of TS. Finally, we acknowledge that this computable phenotype only accounts for patients who are clinically diagnosed with TS and this diagnosis is documented in the EHR. There are undoubtedly patients with TS who have a delayed and/or missed diagnosis, and future studies could use machine learning approaches to identify EHR variables that predict TS in order to identify patients who do not have a billing diagnosis of TS in their EHR (Alexandrou et al., 2020; Berglund et al., 2019; Khalid et al., 2023).

In conclusion, this study successfully developed and validated a computable phenotype for TS in PEDSnet, making it feasible to conduct studies in this population utilizing big data sources. To our knowledge, this is the first computable phenotype validation study for any pediatric genetic condition. The algorithm with the highest C-statistic employed a minimum of three billing diagnosis codes that mapped to TS, although criteria can be adjusted to optimize sensitivity or specificity based on the goals for a particular study. Other variables that may be important in TS research, such as age of TS diagnosis and HRT, were less accurate, and others, such as specific karyotype, age of thelarche or menarche, were unavailable in PEDSnet. These results will inform future TS research seeking to use EHR data, such as comparative effectiveness research, epidemiological trends in diagnosis and management, and prediction modeling to individualize care. These results can also be used when implementing computable phenotyping approaches for other conditions, especially pediatric rare disease and/or genetic disorders.

## Data Availability

The results reported here are based on detailed individual-level patient data compiled as part of the PEDSnet Program. Due to the high risk of reidentification based on unique patterns in the clinical data, even when demographic identifiers have been removed, patient privacy regulations prohibit us from releasing the individual-level data publicly. The data are maintained in a secure enclave, with access managed by the program coordinating center to remain compliant with regulatory and program requirements. Please direct requests to access the data, either for reproduction of the work reported here or for other purposes to.

## Notes

### Competing Interest Statement

SMD and PYF are site investigators for a clinical trial of growth hormone in Turner syndrome sponsored by Ascendis Pharma. NJN is a consultant for Neurocrine Biosciences, Inc. and Ionis Pharmaceuticals. CI is a site investigator for clinical trial of growth hormone in children with growth hormone deficiency and Turner syndrome sponsored by Novo Nordisk, and treatment of type 2 diabetes in children sponsored by Eli Lilly. All other authors have no conflicts of interest to declare.

### Funding Statement

This study was in part funded by NIH/NICHD K23HD092588 and R03HD102773 (SD) and the Doris Duke Foundation (SD, NN). Contents are the authors' sole responsibility and do not necessarily represent view of the funders. The funders had no role in the design and conduct of the study.

### Author Declarations

IRB of Children's Hospital of Philadelphia (CHOP)(#14-011242, 16-012878, and 16-013563) gave ethical approval for this work.

