## Supplemental Tables for "Development and Validation of a Computable Phenotype for Turner Syndrome Utilizing Electronic Health Records from a National Pediatric Network"

### SUPPLEMENTAL INFORMATION

| Supplemental Table 1. Codebook | | |
|---|---|---|
| **Name/Description** | **Vocabulary** | **Code(s)** |
| Turner syndrome | SNOMED CT | 4307885, 46271763, 46271771, 4007429, 4004648, 4007558, 4316871, 201951, 4004647, 46271765, 4007563, 4116329, 4007091, 4111625, 4114977 |
| Estradiol for hormone replacement therapy | ATC | G03CA (all subcodes) |
| ATC = Anatomical Therapeutic Chemical | | |

**Supplemental Table 2. Characteristics of the 14 false positive cases (inaccurately identified Turner syndrome) from primary analysis**

| | 1 | 2 | 3 | 4 | 5 | 6 | 7 | 8 | 9 | 10 | 11 | 12 | 13 | 14 |
|---|---|---|---|---|---|---|---|---|---|---|---|---|---|---|
| Common TS Feature (e.g. short stature, delayed puberty, premature ovarian insufficiency) | ✓ | ✓ | ✓ | ✓ | ✓ | ✓ | ✓ | ✓ | ✓ | ✓ | ✓ | ✓ | ✓ | ✓ |
| Genetic Testing | ✓ | ✓ | ✓ | ✓ | ✓ | ✓ | ✓ | ✓ | ✓ | ✓ | | ✓ | ✓ | ✓ |
| Estradiol Prescription | | ✓ | ✓ | | ✓ | | ✓ | ✓ | ✓ | ✓ | ✓ | | | ✓ |
| Difference in Sex Development diagnosis | | | | | ✓ | | ✓ | ✓ | | | | | | ✓ |
| May-Thurner Syndrome | | | | | | | | | | | ✓ | | | |
| Other Genetic Diagnosis | | | ✓ | ✓ | ✓ | ✓ | ✓ | ✓ | | ✓ | | ✓ | | ✓ |
